# Genomics-informed outbreak investigations of SARS-CoV-2 using civet

**DOI:** 10.1101/2021.12.13.21267267

**Authors:** Áine O’Toole, Verity Hill, Ben Jackson, Rebecca Dewar, Nikita Sahadeo, Rachel Colquhoun, Stefan Rooke, JT McCrone, Martin P McHugh, Sam Nicholls, Radoslaw Poplawski, The COVID-19 Genomics UK (COG-UK) Consortium, COVID-19 Impact Project (Trinidad & Tobago Group), David Aanensen, Matt Holden, Tom Connor, Nick Loman, Ian Goodfellow, Christine V. F. Carrington, Kate Templeton, Andrew Rambaut

## Abstract

The scale of data produced during the SARS-CoV-2 pandemic has been unprecedented, with more than 5 million sequences shared publicly at the time of writing. This wealth of sequence data provides important context for interpreting local outbreaks. However, placing sequences of interest into national and international context is difficult given the size of the global dataset. Often outbreak investigations and genomic surveillance efforts require running similar analyses again and again on the latest dataset and producing reports. We developed civet (cluster investigation and virus epidemiology tool) to aid these routine analyses and facilitate virus outbreak investigation and surveillance. Civet can place sequences of interest in the local context of background diversity, resolving the query into different ’catchments’ and presenting the phylogenetic results alongside metadata in an interactive, distributable report. Civet can be used on a fine scale for clinical outbreak investigation, for local surveillance and cluster discovery, and to routinely summarise the virus diversity circulating on a national level. Civet reports have helped researchers and public health bodies feedback genomic information in the appropriate context within a timeframe that is useful for public health.

## Introduction

The timely sharing of genomic data during the SARS-CoV-2 pandemic has enabled large-scale national and international surveillance efforts around the world. On a finer scale, pathogen genomics can supplement infection prevention and control efforts in clinical settings, as well as aid in outbreak investigations in community settings (Köser 2012, Quick 2014, Houldcroft 2018, Brown 2019). However, the intense SARS-CoV-2 sequencing effort has produced a genomic dataset orders of magnitude larger than any previous epidemic, with more than 5 million sequences shared publicly at time of writing. It is therefore challenging to effectively condense information into relevant summaries and provide meaningful context in a timeframe that allows the data to be of immediate use to those involved in local outbreak response.

Analysing or interpreting genomic information alone without relevant epidemiological information can be misleading and lead to incorrect conclusions due to the incomplete nature of the data. The relatively low mutation rate of SARS-CoV-2, frequent occurrence of convergent mutations (homoplasies), and prevalence of incomplete genome sequences make it critical to integrate epidemiological information alongside the genomic data to provide the most accurate picture and extract the most value from any given dataset. This includes temporal and spatial information, but may also include outbreak-specific data such as profession, ward, clinical metadata, or the background of viral lineages actively circulating in the community. Outbreak investigations often require bespoke reports that present information in a transparent and accessible manner. The data presented must be easily interpretable by health care providers and teams involved in infection control, the majority of whom are not accustomed to incorporating this type of data into their decision making processes.

The virus genomics community has developed a number of tools for analysing and visualising virus genomic data on the order of magnitude of this pandemic. HgPhyloPlace uses UShER to rapidly place sequences of interest into a global SARS-CoV-2 phylogeny (https://hgwdev.gi.ucsc.edu/cgi-bin/hgPhyloPlace; Turakhia et al 2021). Tree visualization tools such as Pando (<u>pando.tools</u>), cov2tree (<u>cov2tree.org</u>), Microreact (Argimón et al 2016) and Dendroscope (Huson et al 2007) can efficiently display phylogenies with a million sequences. However, even with these innovations, it is challenging to construct a phylogenetic tree of that size, given the particular challenges of SARS-CoV-2 data (De Maio et al 2020, Morel et al 2021). NextStrain takes an alternative approach and downsamples the dataset heavily, leaving a manageable amount of data to display (Hadfield et al 2018). The advantage is a rapidly generated phylogeny, however only a small subset of the full diversity is represented. Approaches to condense SARS-CoV-2 genomic information by Single Nucleotide Polymorphism (SNP) typing or lineage typing – such as scorpio (https://github.com/cov-lineages/scorpio), aln2type (https://github.com/connor-lab/aln2type) and pangolin (O’Toole et al 2021) – have been useful but present one dimensional data.

We developed civet (Cluster Investigation and Virus Epidemiology Tool) to address this challenge of integrating metadata while condensing huge quantities of genomic data, and thereby aid SARS-CoV-2 outbreak investigations and surveillance efforts. Civet enables robust phylogenetic analysis to be performed, dynamically querying a large background dataset and generating interactive reports integrating both epidemiological metadata and genomic analysis. Both Public Health Scotland and Public Health England have routinely used civet to inform local outbreaks of SARS-CoV-2 and a number of studies have already been published that have used civet as tool for genomic epidemiology (Li et al 2021, Aggarwal et al 2021, Eales et al 2021, Francis et al 2021).

## Methods

Civet is a Python-based tool with an embedded analysis pipeline implemented in Snakemake (Köster et al 2012). Civet outputs the analysis as a customisable, interactive HTML report. We developed civet as part of the ARTIC Network (artic.network) and COVID-19 Genomics UK (COG-UK) projects and it has been hosted on CLIMB-COVID (Nicholls et al 2021), an isolated partition of the Cloud Infrastructure for Microbial Bioinformatics (CLIMB), since July 2020 (Connor et al 2016). Public health agencies and researchers across the UK use civet routinely to aid SARS-CoV-2 outbreak investigations and generate local surveillance reports.

### Background data

To run civet, the user must minimally provide a sequence alignment and metadata file representing the background diversity of the pathogen of interest. Users on CLIMB-COVID have this data provided by the COG-UK Datapipe (https://github.com/COG-UK/datapipe) although a similarly centralised set up could be applied elsewhere. Civet can also generate the background alignment, metadata file and a SNP summary file from an unaligned fasta sequence, such as the bulk download sequence file available from GISAID (Figure 1). This short pipeline first filters genome sequences based on a minimum length and maximum ambiguity content (%N) cut-off. It then maps against a reference sequence (default is the canonical SARS-CoV-2 reference genome Genbank ID: NC_045512.2, but any reference genome can be supplied) using minimap2 v2.17 (Li 2018). The resulting sam file is converted to fasta format with the 5’ and 3’ untranslated regions (UTRs) masked using gofasta (https://github.com/cov-ert/gofasta). We generate the background metadata file by parsing information from the sequence headers.

**Figure 1:**
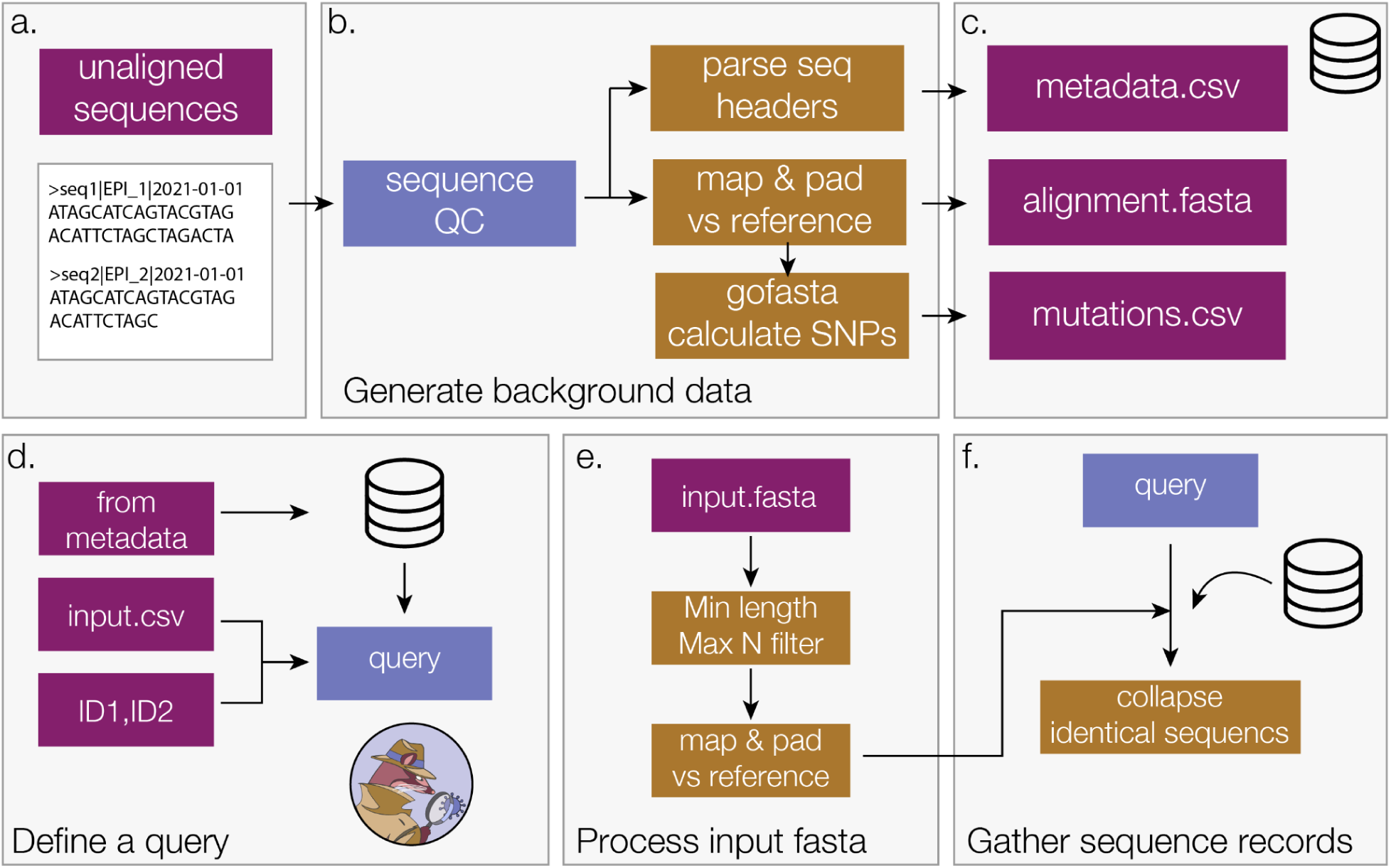
Background data generation pipeline (a-c) and how a civet query is defined (d-f). In order to contextualize the query sequences, civet requires a set of background data files, minimally an alignment and metadata file. a) These files can be generated from an unaligned multi-sequence file using the flag: --generate-civet-background-data. b) The genome sequences are put through a minimum length and maximum N-content filter before being mapped against a reference sequence. The alignment file is generated by trimming and padding against the reference, masking terminal ends with Ns. Information encoded in the sequence header is used to generate the metadata file. gofasta condenses the alignment to the set of derived nucleotide changes in each sequence with respect to the root of the pandemic, to provide an extra speed up for analysis within civet. c) The background files created can then be used as the background data for civet with --datadir or set as an environment variable. d) The query is generated from the background data supplied by specifying a set of criteria to match against, for example all sequences from a particular location within a certain timeframe. The user can also provide a string of specific ids to match or an additional metadata file that specifies the query records and may contain extra metadata fields that only correspond to query sequences, for example patient IDs. e) An additional fasta file for sequences not present in the background data can be provided and civet will perform some quality control checks and align the sequences by mapping and padding against the reference (Default NC_045512.2). f) civet combines the set of query sequence records matched from the background data and from the input fasta file to generate the full query set, and then collapses identical sequences for efficiency. These get expanded out at the end of the analysis pipeline.

### Input options

There are two main ways to define a query dataset, described in Figure 1d. First, a user can define a query from the background data based on metadata, for instance a collection date within a certain time frame, or sequences from a particular location. For example, to generate a report for sequences from June 2021 sampled in Edinburgh: civet --from-metadata date=2021-06-01:2021-07-01 location=Edinburgh. Alternatively, the user can supply a string of query identifiers directly to civet, or a comma-separated (CSV) file specifying the query sequences with some additional metadata not present in the background, like patient IDs. Optionally, a separate fasta file can be supplied to run an analysis on sequences not present in the background dataset. The sequences will go through configurable quality control filters for minimum sequence length and maximum N-content, and are then aligned by mapping and padding against the reference sequence as described for the background dataset creation (Figure 1e). Query identifiers are matched with the alignment in the background data, and the set of fasta sequence records is compiled from queries in both alignment files. Identical sequences are collapsed to a single unique sequence (Figure 1f). Collapsing identical sequences greatly improves analysis efficiency, particularly for outbreak investigations of epidemiologically linked sequences.

### Analysis pipeline

Once identical sequences have been collapsed, civet searches the background dataset using the ‘updown-top-ranking’ method in gofasta v0.0.5 (https://github.com/cov-ert/gofasta) to identify the local set of sequences most similar to each query. Comparing the set of derived SNPs in each query with the set of derived SNPs in every record (target) in the background dataset (Figure 2a) this algorithm can efficiently extract genetically similar genomes from a dataset comprising millions of records. As illustrated in Figure 2a, SNPs can either be unique to the query sequence, unique to the target sequence, or present in the intersection of the two. SNPs present in the intersection represent shared ancestry whereas an excess of SNPs in either the query or target set can be interpreted to give directionality relative to a root sequence. These set comparisons (details in Supplementary Figure 1) allow the target sequences to be classified as either on a polytomy with (same), a direct ancestor of (up), a direct descendant of (down) or polyphyletic with (side) the query sequence (Figure 2). Each target is then ranked according to SNP distance from the query sequence (as illustrated in the schema in Figure 2b). The customisable SNP distance is used to define which target sequences fall within the catchment of a given query. All equally distant targets are included in the catchment. For a given query, if no targets fall within the SNP distance cut off, the algorithm continues outwards in all directions and attempts to get at least one sequence per category (up, down or side). This results in a set of targets for each query, and any queries with overlapping targets have their catchments merged together (Figure 2c).

**Figure 2:**
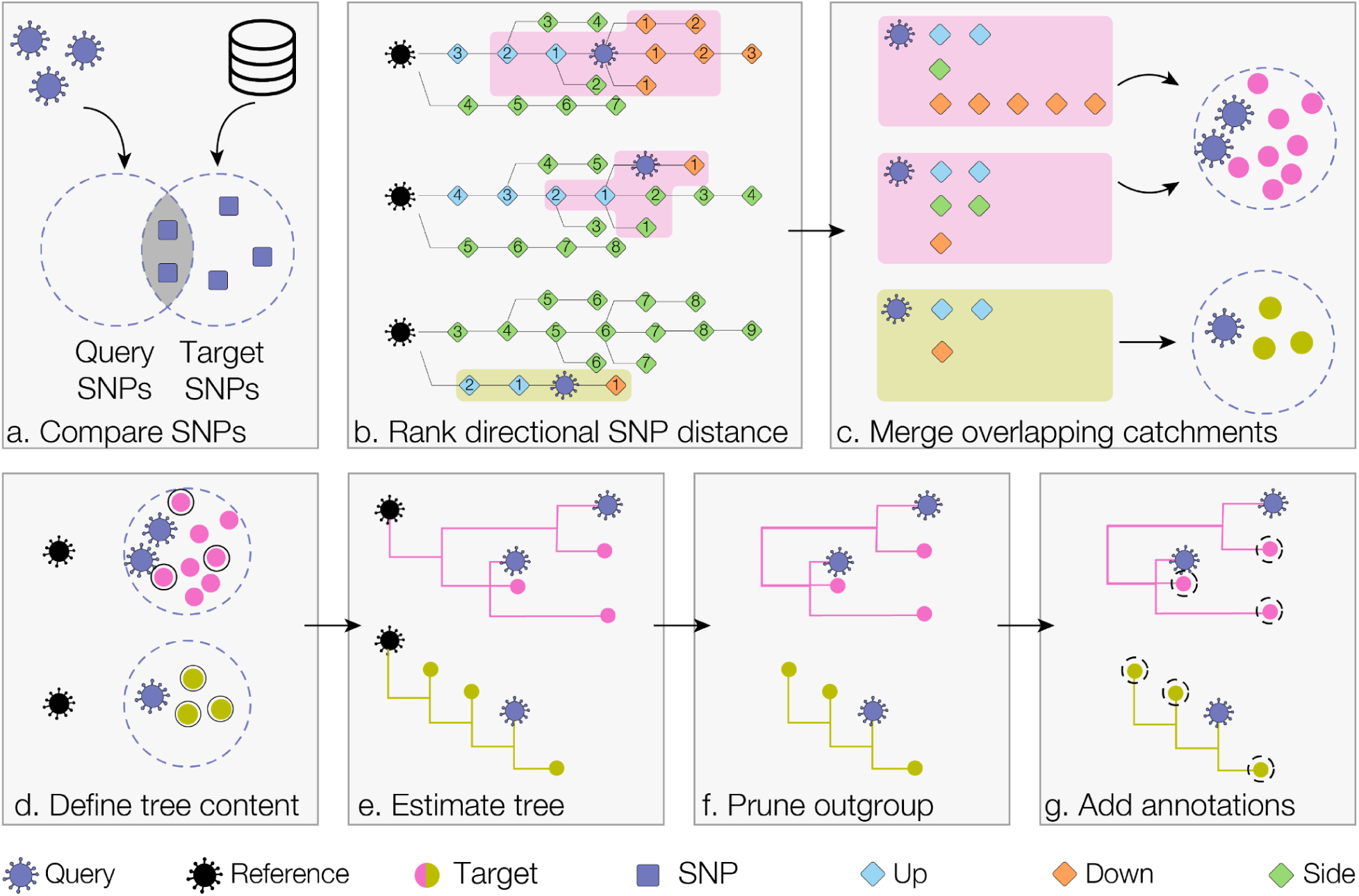
Schema of civet catchment and tree building pipeline. We show three query sequences, falling in two distinct catchments (pink and green). a) Each query sequence is compared against the set of SNPs for every record (target) in the background metadata. By evaluating the intersection and union of the two SNP sets, it is possible to assess directional SNP distance relative to the reference sequence (the early lineage A sequence with GISAID ID EPI_ISL_406801). b) For each query, all targets are ranked by distance from the query and classified as either up, down or side targets based on the set profile in panel a. c) Catchments are constructed by selecting all targets that fall within the specified SNP distance. Up, down and side distances can be configured separately (the default SNP distance of 2 SNPs for all categories is shown here). Civet then merges any catchments with overlapping targets. d) An outgroup reference sequence is added to each catchment and, if necessary, catchments are downsampled. e) Civet estimates a maximum likelihood tree for each catchment using iqtree. f) The reference sequence is pruned out and the tips of the tree are annotated with user-specified fields. g) Specific metadata annotations are added to each tip, which can be toggled within the report.

At this point, there is no limit to the size of catchments and as the pandemic has been sampled so intensively in some areas, even relatively low SNP distances can lead to a large catchment. The user has the option to downsample the catchments prior to tree building and configure the maximum number of the background sequences to include in a given catchment tree (Figure 2d). Downsampling can be run in: random mode, which randomly samples from the full catchment; enrich mode, which allows the user to specify a metadata trait to enrich for and the factor by which to enrich over the other targets in the catchment; or normalise mode, which allows the user to sample evenly across a metadata trait, such as epiweek. The query sequences, background catchment sequences and an anonymised early lineage A outgroup sequence are then gathered for tree building. Each catchment tree along with the queries is then estimated using iqtree with the HKY substitution model, in fast mode (Minh et al 2020, Hasegawa et al 1985; Figure 2e). The civet software then prunes the outgroup from the resulting maximum likelihood trees and annotates them with user-specified metadata traits (Figure 2f-g). Optionally, the user can search for mutations of interest and investigate which nucleotide or amino acid variant is present at sites in both the queries and background catchment sequences, and can also annotate these in the catchment trees.

### Report content

Civet generates a fully customisable report, summarising information about the queries of interest and the surrounding diversity. The report generated is a HTML file that can be viewed in a web browser, thus allowing the interactivity of web-pages. The components of the report include an interactive table summarising metadata of the query sequences, including any user supplied metadata; which catchment a query falls in; and the mutations of interest if specified. This table can be sorted, filtered and its columns can be dynamically configured, all within the distributable report. For each catchment, the civet report contains a table summarising the catchment content (prior to downsampling) and describes which lineages and countries are present in this local diversity neighbourhood (example shown in Figure 4b).

The civet report displays the catchment trees using the interactive tree visualisation library FigTree.js (https://github.com/rambaut/figtree.js). The trees can be expanded out along the vertical axis and tip nodes can be coloured by any field specified with annotations --tree-annotations. Clades can be collapsed down by clicking on the parent branch and uncollapsed by clicking again. Each taxa in the tree is associated with additional metadata that can be displayed by selecting a tip (demonstrated in Figure 4d). Civet runs snipit, a python tool that finds the SNPs relative to a reference in a multiple sequence alignment and highlights these changes as a figure (https://github.com/aineniamh/snipit). The report also contains a query timeline based on supplied temporal metadata, and interactive maps both for plotting the query sequence locations and for summarising the background diversity in the location of interest up to administrative level 2 for the UK and administrative level 1 for the rest of the world.

The user can generate multiple reports with one command to customise content for different intended audiences. Using the --report-content option, a report containing all the results shown in Figure 4 can be generated alongside a report intended for the Infection Prevention and Control (IPC) team, which may just contain the summary tables for instance and not the phylogenies Full report configuration details can be found at the civet documentation at https://cov-lineages.org/resources/civet.html.

## Results

### Hospital outbreak

There have been a number of studies demonstrating the utility of in-hospital genomic epidemiology for outbreak investigation to supplement standard infection prevention and control (IPC) practices (e.g. Houldcroft et al 2018, Brown et al 2019, Stirrup 2021). To aid in these investigations, which generally involve standard bioinformatic and phylogenetic methods and report generation, civet can contextualize sequences of interest and generate distributable routine reports.

The case study presented in Figure 3 describes an outbreak investigation carried out in an Edinburgh hospital in 2020. An outbreak of SARS-Cov-2 was detected, with cases across three wards that included multiple staff and patients (Figure 3a). The earliest case detected was a patient in Ward B sampled on Day 0 (Figure 3b). In the following days, three more patients across Wards A and B tested positive for SARS-CoV-2, two of whom had recently travelled from Country X. Subsequently, three healthcare workers who had been working in Ward A and two healthcare workers who had been working across Wards B and C tested positive. A household contact of one of these healthcare workers tested positive the same day and finally a healthcare worker in ward C tested positive. At the outset of the investigation, the outbreak was thought to have been caused by either an initial patient to staff transmission event with subsequent staff to staff transmission, or multiple patient to staff exposures.

**Figure 3.**
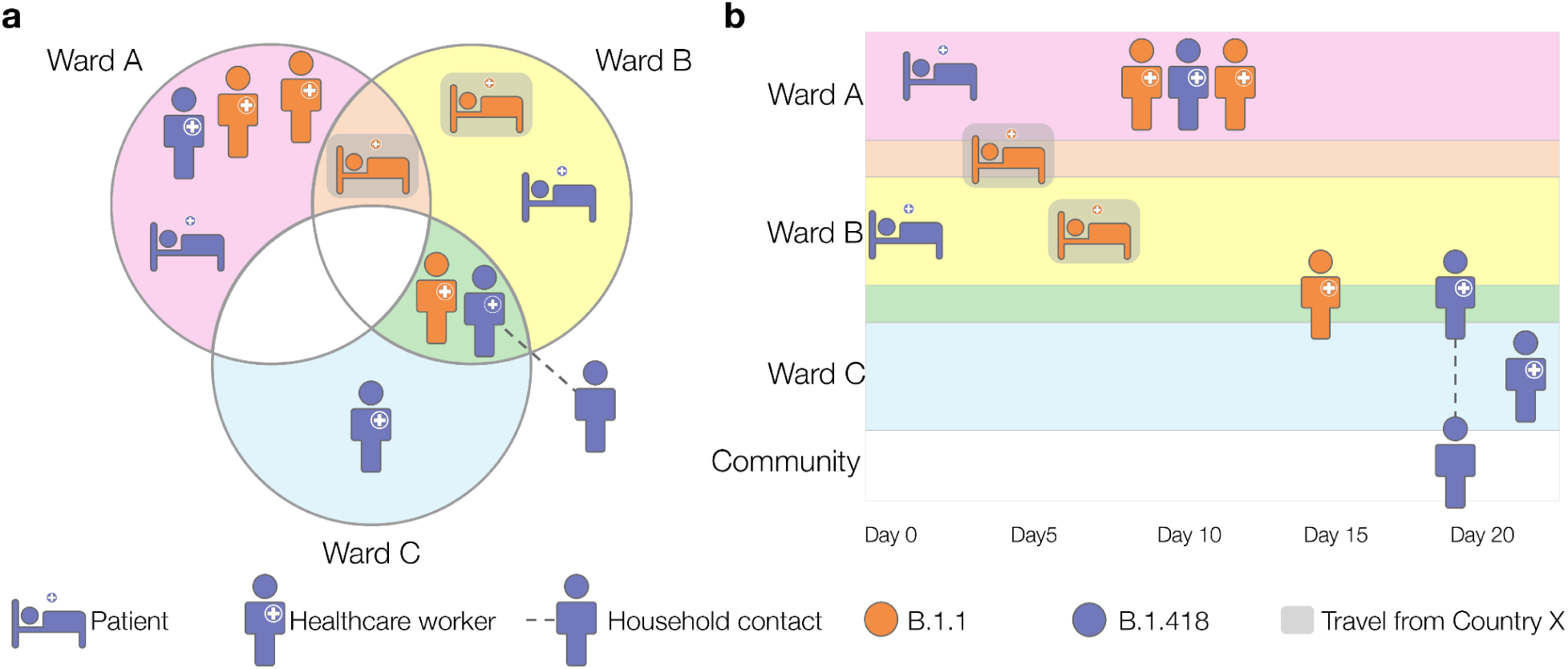
Schema of clinical outbreak investigation June 2020, colour of cases indicate lineage revealed by genome sequencing (B.1.1 or B.1.418). a) The outbreak occurred across three wards and involved six members of staff, four patients and one household contact of a staff member. b) Timeline of sample collection dates across wards A, B and C.

**Figure 4.**
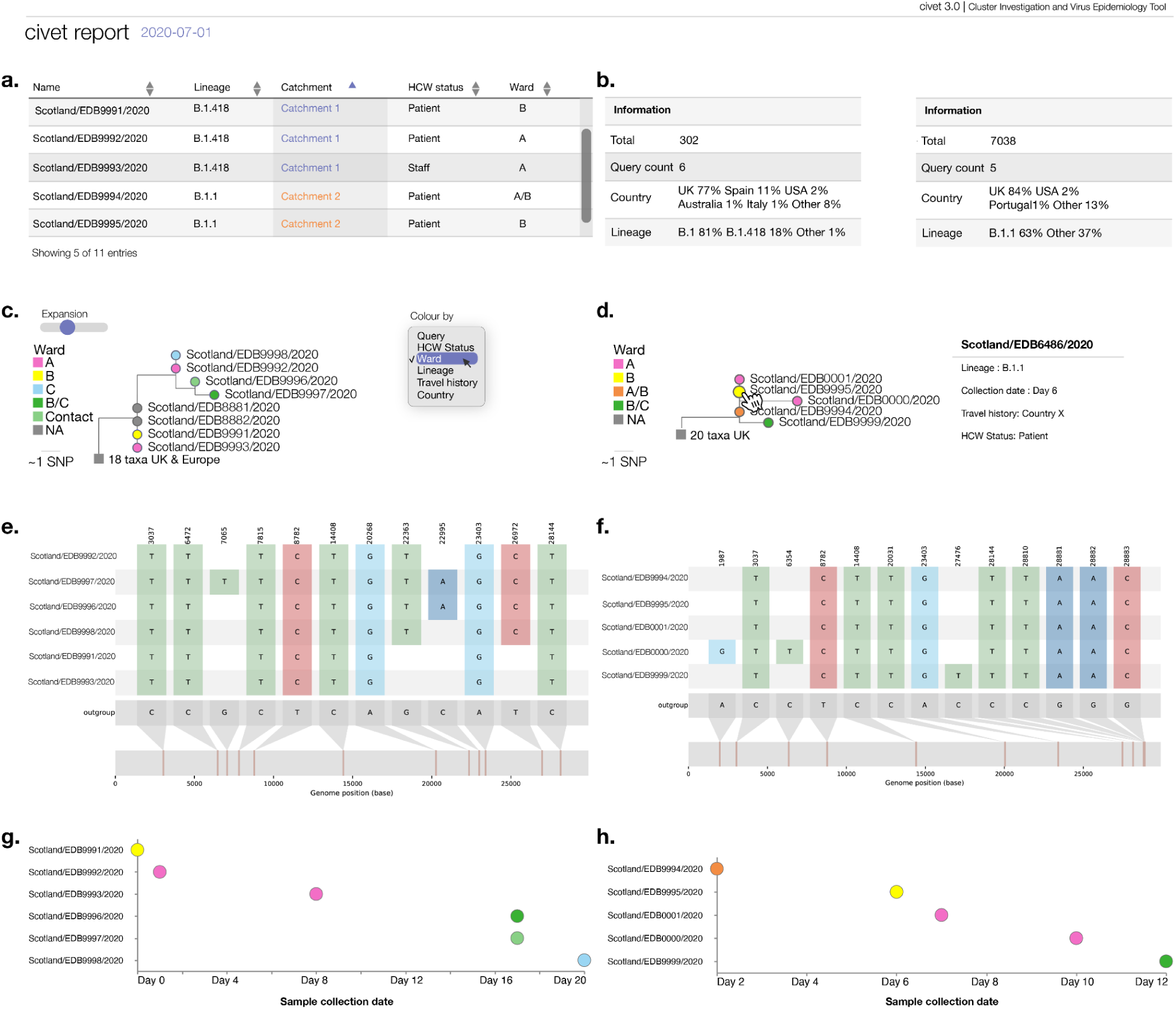
Components of a civet report generated for the outbreak investigation in a clinical setting described in Figure 3. a) The metadata of all query sequences is summarised in an interactive table, with sortable columns that can be toggled on and off. b) Each catchment is summarised in full, regardless of downsampling. Number of queries and the countries and lineages within the catchment are indicated. c) The catchment phylogenies are displayed initially in compact form, but can be expanded vertically using the Expansion slider. By default tip nodes are coloured by whether a tip is a query taxa or not, but the dropdown menu allows the user to colour tip nodes by any trait specified in --tree-annotations. d) Tip nodes can be selected to show the metadata associated with that particular sequence and clades can be collapsed to a single node by selecting the parent branch. e-f) snipit graphs highlight nucleotide differences from the reference genome. g-h) A timeline summarises any query date information provided. Note: all metadata has been de-identified for data protection purposes.

Genome sequencing of SARS-CoV-2 samples from staff and patients revealed the outbreak consisted of two distinct clusters, or catchments, corresponding to PANGO lineages B.1.1 and B.1.418. Figure 4 summarises the content of the default report produced by civet, full report available at https://cov-lineages.org/resources/civet/civet_case_study_1.html. Figure 4a displays the interactive query summary table and catchment summary tables (Figure 4b). The phylogenies in Figure 4c and 5d are coloured by ward. Figure 4c shows the phylogenetic relationship of queries present in catchment 1, alongside the background sequences. Two community samples also from Edinburgh sit on a polytomy with, and are identical to, the earliest patient case detected in Ward B. Particularly with SARS-CoV-2 it’s not possible to infer directionality based on this information, however this phylogeny does show that the diversity in the hospital overlapped with that present in the community. Figure 4d shows the phylogenetic relationship of catchment 2, with the two patients with travel history from Country X and earliest staff member to contract lineage B.1.1 all sharing identical SARS-CoV-2 genome sequences. Figure 4e displays the snipit plots that summarise the nucleotide changes from reference among queries of interest, and the sample collection date for each query sequence is shown in the timeline plot in Figure 4f, coloured by ward. civet resolved the outbreak into two distinct catchment trees making it likely that there were multiple introductions into the hospital from the community, and the mixture of wards present in each catchment implies some between-ward transmission. This report highlights two areas of control for the IPC to focus future efforts. As the case was deemed at least two separate introduction events with clear transmission links, the outbreak investigation was subsequently closed by the IPC team.

**Figure 5:**
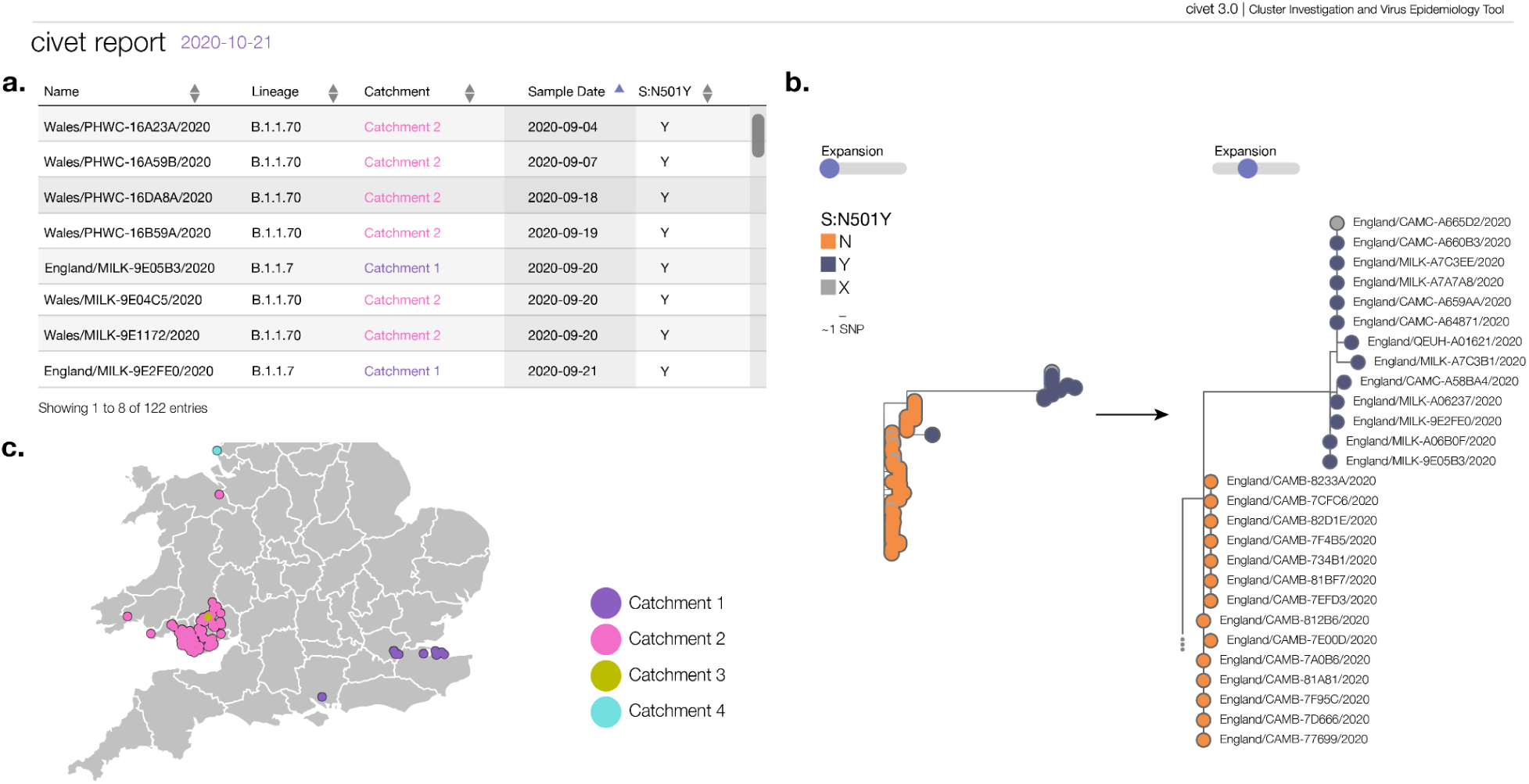
Sample of figures from a civet report demonstrating its use for community surveillance in the UK. As a hypothetical example, we used civet to search the COG-UK dataset from the 21st of October 2020 for SARS-CoV-2 sequences with the spike protein mutation N501Y in September and October 2020. At this point, 4 independent occurrences of this mutation were detected using civet. The earliest sequences can be seen in panel a. The two main clusters correspond to B.1.1.70, which was a lineage circulating in Wales, and B.1.1.7, which only had 13 sequences at this time point. Despite being small, the striking basal branch of B.1.1.7 is clearly visible in panel b. Running civet routinely enables early identification and tracking of clusters such as these. Panel c shows the query map of the samples identified with N501Y and the geographic separation of catchments 1, 2 and 4.

### Community surveillance

Civet can also be used as part of routine local surveillance to summarise the diversity of viruses circulating in a local area or to flag and monitor clusters of interest. The N501Y mutation in the SARS-CoV-2 spike protein has been predicted to increase SARS-CoV-2 receptor binding domain ACE2 affinity (Starr et al 2020; https://jbloomlab.github.io/SARS-CoV-2-RBD_DMS/ last accessed 2021-08-10). As such, the presence of this mutation has been monitored as part of the genomic surveillance efforts in the UK and around the world. We present a hypothetical case of a civet report generated from a simple command used to search a background dataset from COG-UK from the 21st of October 2020 (Figure 5, full report available at https://cov-lineages.org/resources/civet/civet_case_study_2.html). The search defined queries as sequences from the UK with the spike N501Y mutation from the beginning of September 2020 to the latest data in the background set (2020-10-21). Figure 5a demonstrates the query summary table sorted by earliest samples. At the time in the UK, two concurrent geographically-distinct clusters existed (Figure 5c); one in Wales that became known as B.1.1.70 and one in south east England that became B.1.1.7. There were also two further, very small, clusters that contained S:N501Y between 1st September and 21st October 2020. At this snapshot in time, B.1.1.7 is clearly distinguishable but only has 13 sequences. By running civet routinely, the user can both discover and monitor clusters such as B.1.1.7 and B.1.1.70 as they progress, facilitating rapid public health interventions.

### National surveillance

Civet also has the flexibility to inform surveillance efforts at the national level. In Figure 6, we show a schema of a civet report summarising genomic surveillance efforts in Trinidad and Tobago during 2020, full report available at https://cov-lineages.org/resources/civet/civet_case_study_3.html. Figure 6a displays the Trinidad and Tobago sequences alongside the available metadata, and summarises how many distinct catchments the genomes are represented by. Sequences from Trinidad and Tobago fall within three catchments, which correspond to lineages B.1.111, B.1.1 and B.1.1.33. The presence of three distinct catchments indicates there were at least three independent introductions into Trinidad and Tobago during 2020. Figure 6b and 6c show the phylogeny for catchment 1. The Trinidad and Tobago sequences form a monophyletic cluster within the background diversity of sequences from countries around the world. The timeline of events can be seen in Figure 6d, with lineage B.1.111 appearing throughout the latter half of 2020, and B.1.1 and B.1.1.33 appearing only transiently. We summarise the background diversity of other nations with SARS-CoV-2 genome data from 2020 on public databases in Figure 6e. Trinidad and Tobago is highlighted with a schema of the tooltip available in the interactive civet report. This report gives a picture of how Trinidad and Tobago fits into the overall diversity of SARS-CoV-2 in 2020. Reports could be routinely generated on a weekly or monthly basis to provide information on the changing context of a country’s epidemic compared to its neighbours.

**Figure 6:**
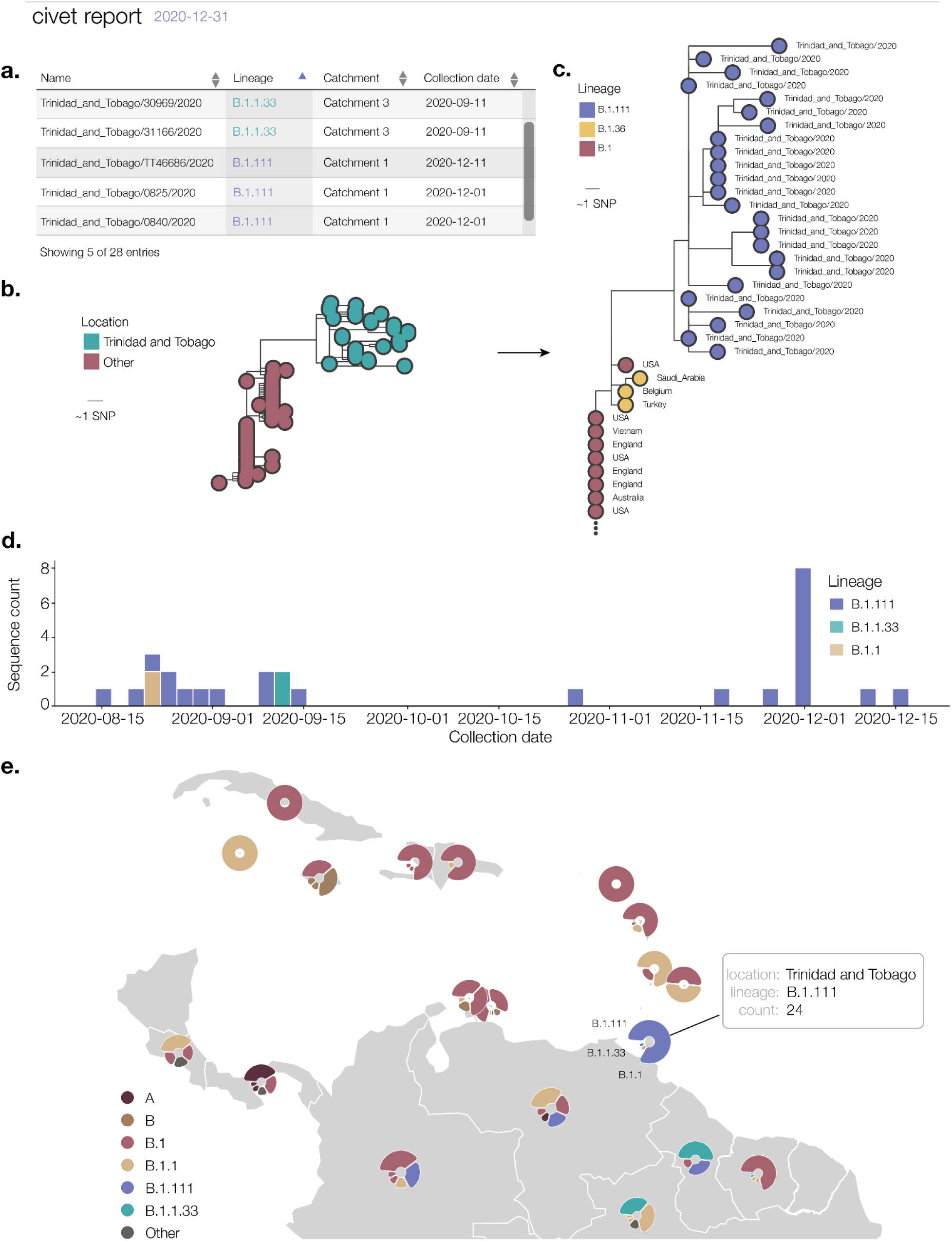
Schema of a national level surveillance report generated using civet for Trinidad and Tobago. All SARS-CoV-2 genome sequences on GISAID from 2020 with <20% ambiguity content are summarised in the report (n=28). a. Available metadata for query sequences from Trinidad and Tobago. Most genomes have been assigned lineage B.1.111, although a smaller number of genomes are assigned other lineages B.1.1.33 and B.1.1. b. Catchment 1 phylogeny. Query sequences are placed in the context of background diversity beyond Trinidad and Tobago. c. Expanding the phylogeny and colouring tips by lineage shows this catchment includes query sequences from lineage B.1.111. d. Aggregate count of queries over time, coloured by lineage. e. Lineage diversity of Trinidad and Tobago and surrounding countries as generated using the background diversity map in civet.

## Discussion

Virus genome sequencing can help reveal transmission chains and clusters of interest to aid outbreak investigations and surveillance efforts, as exemplified by the case studies above. With civet, academic researchers and public health scientists can easily run complex and robust phylogenetic analyses with a single command, contextualising sequences of interest in the large background dataset and visualising them alongside temporal, spatial and other epidemiological metadata in an interactive, distributable report. This frees users to place emphasis on interpreting the data and allows them to deliver information on a time-frame that is useful for public health responses.

Throughout the SARS-CoV-2 pandemic, civet has been primed for use investigating SARS-CoV-2 clinical outbreaks and running local surveillance on CLIMB-COVID (Nicholls et al 2021) as part of the COG-UK project. Each day on CLIMB-COVID, researchers from around the UK upload the latest SARS-CoV-2 genome sequences and accompanying metadata. The read data undergo rigorous quality control and a data-processing and phylogenetics pipeline compiles and analyzes the resulting genomes in combination with the global dataset from GISAID (https://github.com/COG-UK/datapipe). This makes the latest SARS-CoV-2 genome data available to civet users on a daily basis. COG-UK data protection stipulates that data cannot be removed from CLIMB-COVID and often outbreak investigations involve sensitive, protected metadata. With civet, researchers can run analysis on CLIMB-COVID, distribute the report and keep their metadata protected. Civet has been popular and widely used within the framework of COG-UK, by academic researchers and scientists in public health agencies, for investigating SARS-CoV-2 clinical outbreaks and running local surveillance. A similar centralised server infrastructure could be set up for a national surveillance response or more local “locked down” compute environments (Nicholls et al 2021) and civet could be easily implemented within this framework to aid outbreak investigations.

Civet can easily perform phylogenetic analysis on large datasets and provide reports for any countries with sequences to analyse. Default settings are configured for SARS-CoV-2, but civet is virus-agnostic and can be set up to run on other viruses of interest with an appropriate background dataset and reference sequence. Although civet is currently a command-line based tool, a clear extension to the software is to develop and provide a graphical user interface. This will enable users unfamiliar with the command line to run civet. We also plan to continue developing civet and adding extra features, including a country specific summary comparing counts of genomes sequenced over time with additional epidemiological data such as cases per country over time, which is already available on the Johns Hopkins University COVID-19 DataAPI (Dong et al 2020). This particular feature will help give appropriate context for countries with relatively low numbers of sequences as it is important to keep sequencing biases into account when inferring outbreak or transmission dynamics.

As the ability to rapidly sequence pathogens at scale has become less technically challenging, in part due to the availability of robust protocols such as those by the ARTIC Network (Quick 2017), the amount of data that can be generated from a small laboratory with limited infrastructure has significantly increased. Arguably the greatest challenges now lay at trying to best utilise this data in an effective way to inform the response efforts, which hinges entirely on the ability to efficiently contextualise the data and provide an output that is interpretable by those less versed in the interpretation of phylogenetic trees. In this way, civet can help alleviate the analytical bottleneck that exists as a major issue for many public health labs and can maximise the value of genomic data.

## Data Availability

All code presented is available on GitHub.

https://github.com/artic-network/civet

## Acknowledgements

We thank the following for helpful suggestions, comments, beta-testing, feature requests and patience: Matt Loose, Matt Bashton, Richard Myers, Meera Chand, Anthony Underwood, Ben Lindsey, Jeff Barrett, Derek Fairley, Joseph Hughes, David Robertson, Richard Orton, Ulf Schaefer, Natalie Groves, Nikos Manesis, Jayna Raghwani. We acknowledge the hard work and ethos of open-science of the individual research labs and public health bodies that have made their genome data accessible on GISAID.

## Availability of data and materials

Project home page: https://github.com/artic-network/civet

Operating system(s): Unix based platforms, tested on Ubuntu and MacOSX

Programming language: Python, mako

All code is open-source and available on GitHub at github.com/artic-network/civet under a GNU General Public License v3.0.

---

**Figure S1.**
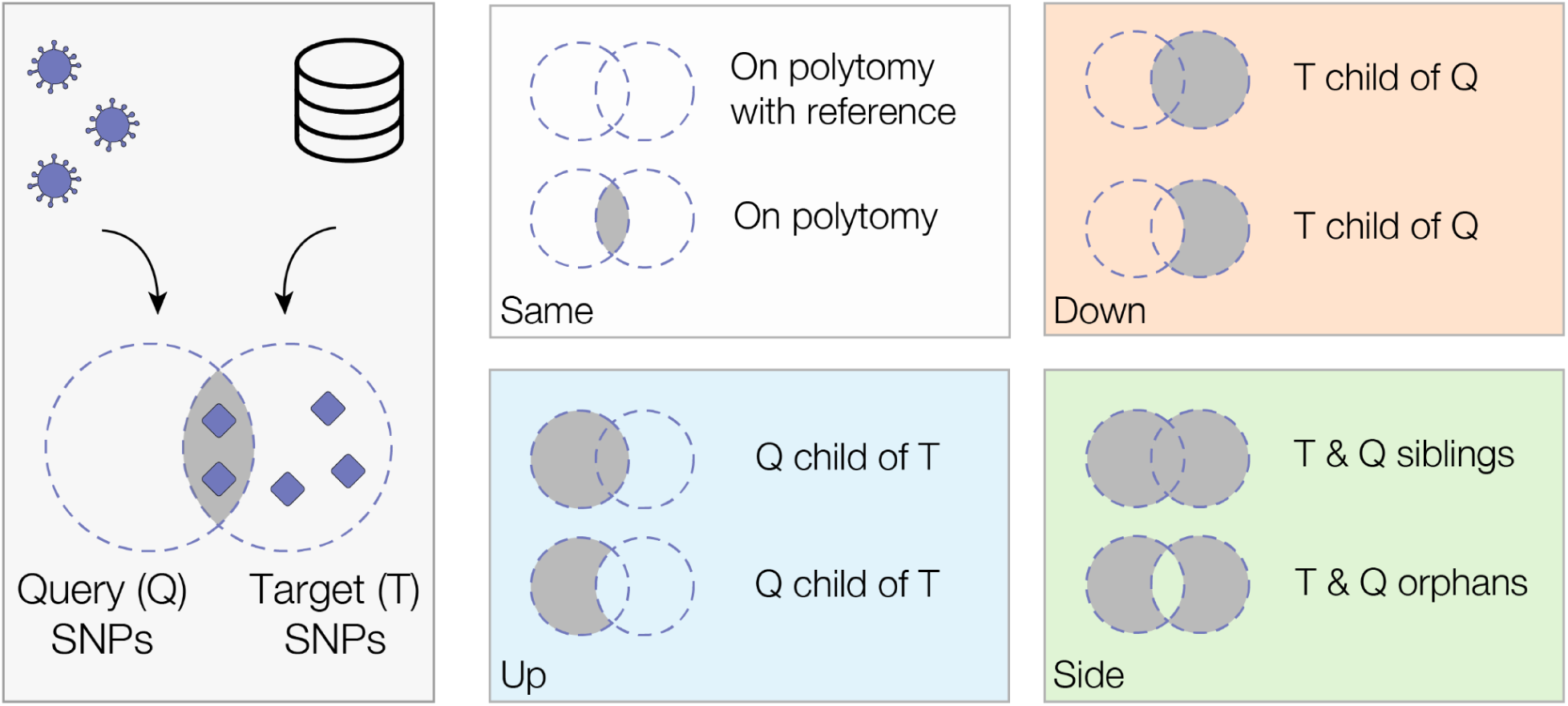
Set categories for gofasta “updown-top-ranking”. Shaded regions in the Venn diagrams represent having at least one SNP in that category (either in Q, T or Q ∩ T).

**Table S1.**
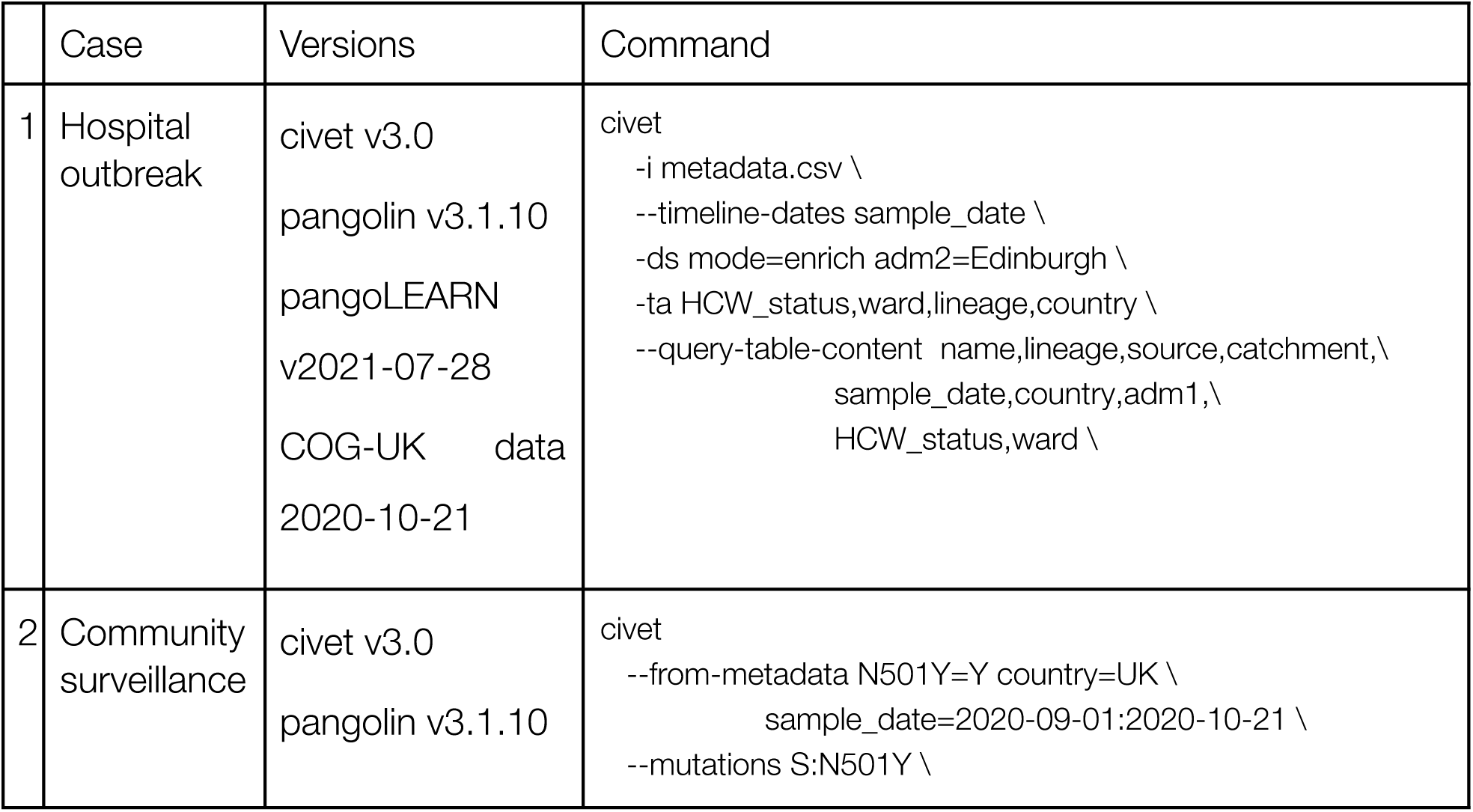
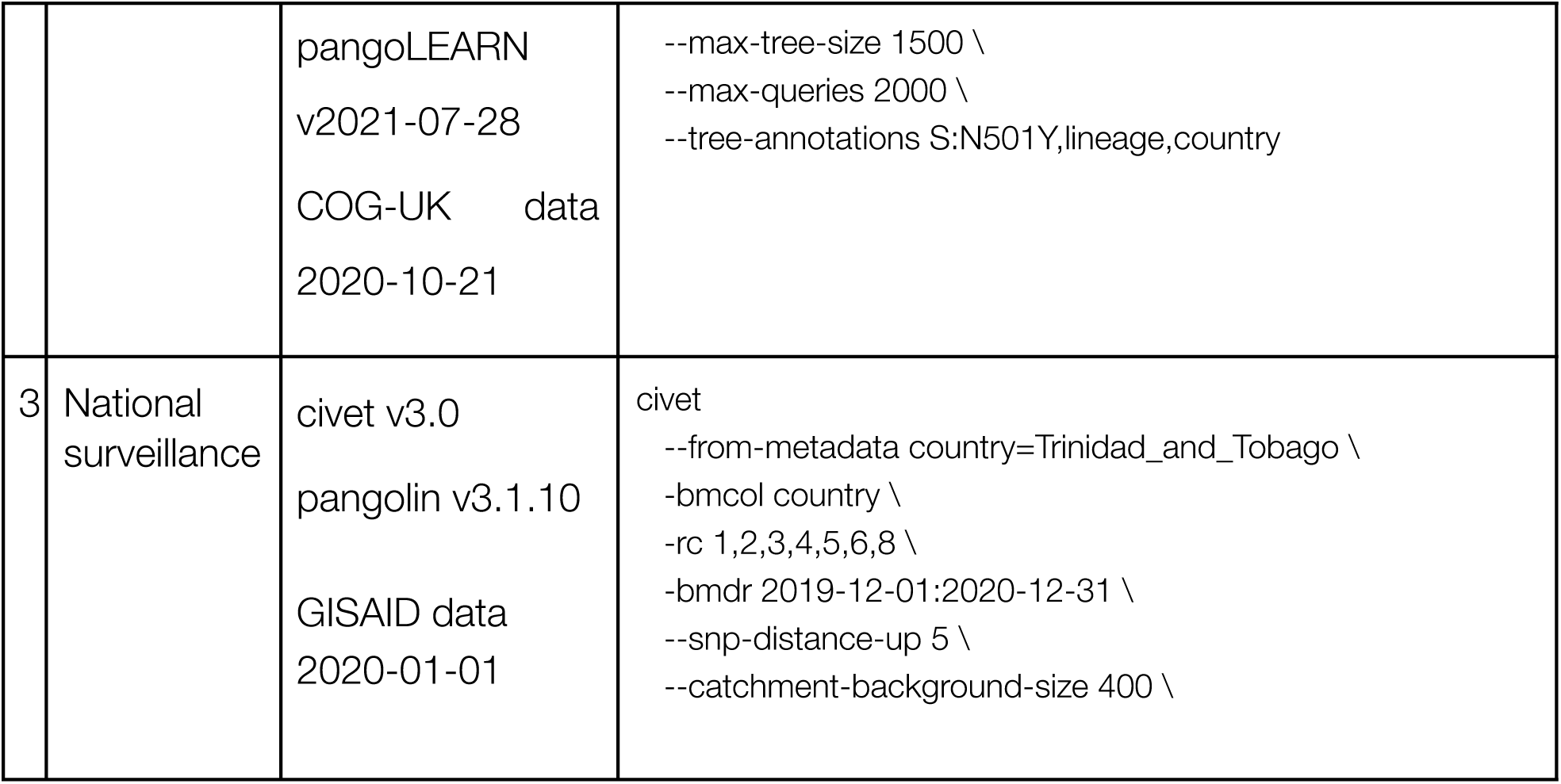
Commands index

## Supplemental author list

COG-UK

Funding acquisition, Leadership and supervision, Metadata curation, Project administration, Samples and logistics, Sequencing and analysis, Software and analysis tools, and Visualisation:

Dr Samuel C Robson PhD ^13, 84^

Funding acquisition, Leadership and supervision, Metadata curation, Project administration, Samples and logistics, Sequencing and analysis, and Software and analysis tools:

Dr Thomas R Connor PhD ^11, 74^ and Prof Nicholas J Loman PhD ^43^

Leadership and supervision, Metadata curation, Project administration, Samples and logistics, Sequencing and analysis, Software and analysis tools, and Visualisation:

Dr Tanya Golubchik PhD ^5^

Funding acquisition, Leadership and supervision, Metadata curation, Samples and logistics, Sequencing and analysis, and Visualisation:

Dr Rocio T Martinez Nunez PhD ^46^

Funding acquisition, Leadership and supervision, Project administration, Samples and logistics, Sequencing and analysis, and Software and analysis tools:

Dr David Bonsall PhD ^5^

Funding acquisition, Leadership and supervision, Project administration, Sequencing and analysis, Software and analysis tools, and Visualisation:

Prof Andrew Rambaut DPhil ^104^

Funding acquisition, Metadata curation, Project administration, Samples and logistics, Sequencing and analysis, and Software and analysis tools:

Dr Luke B Snell MSc, MBBS ^12^

Leadership and supervision, Metadata curation, Project administration, Samples and logistics, Software and analysis tools, and Visualisation:

Rich Livett MSc ^116^

Funding acquisition, Leadership and supervision, Metadata curation, Project administration, and Samples and logistics:

Dr Catherine Ludden PhD ^20, 70^

Funding acquisition, Leadership and supervision, Metadata curation, Samples and logistics, and Sequencing and analysis:

Dr Sally Corden PhD ^74^ and Dr Eleni Nastouli FRCPath ^96, 95, 30^

Funding acquisition, Leadership and supervision, Metadata curation, Sequencing and analysis, and Software and analysis tools:

Dr Gaia Nebbia PhD, FRCPath ^12^

Funding acquisition, Leadership and supervision, Project administration, Samples and logistics, and Sequencing and analysis:

Ian Johnston BSc ^116^

Leadership and supervision, Metadata curation, Project administration, Samples and logistics, and Sequencing and analysis:

Prof Katrina Lythgoe PhD ^5^, Dr M. Estee Torok FRCP ^19, 20^ and Prof Ian G Goodfellow PhD ^24^ Leadership and supervision, Metadata curation, Project administration, Samples and logistics, and Visualisation:

Dr Jacqui A Prieto PhD ^97, 82^ and Dr Kordo Saeed MD, FRCPath ^97, 83^

Leadership and supervision, Metadata curation, Project administration, Sequencing and analysis, and Software and analysis tools:

Dr David K Jackson PhD ^116^

Leadership and supervision, Metadata curation, Samples and logistics, Sequencing and analysis, and Visualisation:

Dr Catherine Houlihan PhD ^96, 94^

Leadership and supervision, Metadata curation, Sequencing and analysis, Software and analysis tools, and Visualisation:

Dr Dan Frampton PhD ^94, 95^

Metadata curation, Project administration, Samples and logistics, Sequencing and analysis, and Software and analysis tools:

Dr William L Hamilton PhD ^19^ and Dr Adam A Witney PhD ^41^

Funding acquisition, Samples and logistics, Sequencing and analysis, and Visualisation:

Dr Giselda Bucca PhD ^101^

Funding acquisition, Leadership and supervision, Metadata curation, and Project administration:

Dr Cassie F Pope PhD^40, 41^

Funding acquisition, Leadership and supervision, Metadata curation, and Samples and logistics:

Dr Catherine Moore PhD ^74^

Funding acquisition, Leadership and supervision, Metadata curation, and Sequencing and analysis:

Prof Emma C Thomson PhD, FRCP ^53^

Funding acquisition, Leadership and supervision, Project administration, and Samples and logistics:

Dr Ewan M Harrison PhD ^116, 102^

Funding acquisition, Leadership and supervision, Sequencing and analysis, and Visualisation:

Prof Colin P Smith PhD ^101^

Leadership and supervision, Metadata curation, Project administration, and Sequencing and analysis:

Fiona Rogan BSc ^77^

Leadership and supervision, Metadata curation, Project administration, and Samples and logistics:

Shaun M Beckwith MSc ^6^, Abigail Murray Degree ^6^, Dawn Singleton HNC ^6^, Dr Kirstine Eastick PhD, FRCPath ^37^, Dr Liz A Sheridan PhD ^98^, Paul Randell MSc, PgD ^99^, Dr Leigh M Jackson PhD^105^, Dr Cristina V Ariani PhD ^116^ and Dr Sónia Gonçalves PhD ^116^

Leadership and supervision, Metadata curation, Samples and logistics, and Sequencing and analysis:

Dr Derek J Fairley PhD ^3, 77^, Prof Matthew W Loose PhD ^18^ and Joanne Watkins MSc ^74^

Leadership and supervision, Metadata curation, Samples and logistics, and Visualisation:

Dr Samuel Moses MD ^25, 106^

Leadership and supervision, Metadata curation, Sequencing and analysis, and Software and analysis tools:

Dr Sam Nicholls PhD ^43^, Dr Matthew Bull PhD ^74^ and Dr Roberto Amato PhD ^116^

Leadership and supervision, Project administration, Samples and logistics, and Sequencing and analysis:

Prof Darren L Smith PhD ^36, 65, 66^

Leadership and supervision, Sequencing and analysis, Software and analysis tools, and Visualisation:

Prof David M Aanensen PhD ^14, 116^ and Dr Jeffrey C Barrett PhD ^116^

Metadata curation, Project administration, Samples and logistics, and Sequencing and analysis:

Dr Dinesh Aggarwal MRCP^20, 116, 70^, Dr James G Shepherd MBCHB, MRCP ^53^, Dr Martin D Curran PhD ^71^ and Dr Surendra Parmar PhD ^71^

Metadata curation, Project administration, Sequencing and analysis, and Software and analysis tools:

Dr Matthew D Parker PhD ^109^

Metadata curation, Samples and logistics, Sequencing and analysis, and Software and analysis tools:

Dr Catryn Williams PhD ^74^

Metadata curation, Samples and logistics, Sequencing and analysis, and Visualisation: Dr Sharon Glaysher PhD ^68^

Metadata curation, Sequencing and analysis, Software and analysis tools, and Visualisation:

Dr Anthony P Underwood PhD ^14, 116^, Dr Matthew Bashton PhD ^36, 65^, Dr Nicole Pacchiarini PhD ^74^, Dr Katie F Loveson PhD ^84^ and Matthew Byott MSc ^95, 96^

Project administration, Sequencing and analysis, Software and analysis tools, and Visualisation:

Dr Alessandro M Carabelli PhD ^20^

Funding acquisition, Leadership and supervision, and Metadata curation:

Dr Kate E Templeton PhD ^56, 104^

Funding acquisition, Leadership and supervision, and Project administration:

Dr Thushan I de Silva PhD ^109^, Dr Dennis Wang PhD ^109^, Dr Cordelia F Langford PhD ^116^ and John Sillitoe BEng ^116^

Funding acquisition, Leadership and supervision, and Samples and logistics:

Prof Rory N Gunson PhD, FRCPath ^55^

Funding acquisition, Leadership and supervision, and Sequencing and analysis:

Dr Simon Cottrell PhD ^74^, Dr Justin O’Grady PhD ^75, 103^ and Prof Dominic Kwiatkowski PhD ^116, 108^

Leadership and supervision, Metadata curation, and Project administration:

Dr Patrick J Lillie PhD, FRCP ^37^

Leadership and supervision, Metadata curation, and Samples and logistics:

Dr Nicholas Cortes MBCHB ^33^, Dr Nathan Moore MBCHB ^33^, Dr Claire Thomas DPhil ^33^, Phillipa J Burns MSc, DipRCPath ^37^, Dr Tabitha W Mahungu FRCPath ^80^ and Steven Liggett BSc ^86^

Leadership and supervision, Metadata curation, and Sequencing and analysis:

Angela H Beckett MSc ^13, 81^ and Prof Matthew TG Holden PhD ^73^

Leadership and supervision, Project administration, and Samples and logistics:

Dr Lisa J Levett PhD ^34^, Dr Husam Osman PhD ^70, 35^ and Dr Mohammed O Hassan-Ibrahim PhD, FRCPath ^99^

Leadership and supervision, Project administration, and Sequencing and analysis:

Dr David A Simpson PhD ^77^

Leadership and supervision, Samples and logistics, and Sequencing and analysis:

Dr Meera Chand PhD ^72^, Prof Ravi K Gupta PhD ^102^, Prof Alistair C Darby PhD ^107^ and Prof Steve Paterson PhD ^107^

Leadership and supervision, Sequencing and analysis, and Software and analysis tools:

Prof Oliver G Pybus DPhil ^23^, Dr Erik M Volz PhD ^39^, Prof Daniela de Angelis PhD ^52^, Prof David L Robertson PhD ^53^, Dr Andrew J Page PhD ^75^ and Dr Inigo Martincorena PhD ^116^

Leadership and supervision, Sequencing and analysis, and Visualisation:

Dr Louise Aigrain PhD ^116^ and Dr Andrew R Bassett PhD ^116^

Metadata curation, Project administration, and Samples and logistics:

Dr Nick Wong DPhil, MRCP, FRCPath ^50^, Dr Yusri Taha MD, PhD ^89^, Michelle J Erkiert BA ^99^ and Dr Michael H Spencer Chapman MBBS ^116, 102^

Metadata curation, Project administration, and Sequencing and analysis:

Dr Rebecca Dewar PhD ^56^ and Martin P McHugh MSc ^56, 111^

Metadata curation, Project administration, and Software and analysis tools:

Siddharth Mookerjee MPH ^38, 57^

Metadata curation, Project administration, and Visualisation:

Stephen Aplin ^97^, Matthew Harvey ^97^, Thea Sass ^97^, Dr Helen Umpleby FRCP ^97^ and Helen Wheeler^97^

Metadata curation, Samples and logistics, and Sequencing and analysis:

Dr James P McKenna PhD ^3^, Dr Ben Warne MRCP ^9^, Joshua F Taylor MSc ^22^, Yasmin Chaudhry BSc ^24^, Rhys Izuagbe ^24^, Dr Aminu S Jahun PhD ^24^, Dr Gregory R Young PhD ^36, 65^, Dr Claire McMurray PhD ^43^, Dr Clare M McCann PhD ^65, 66^, Dr Andrew Nelson PhD ^65, 66^ and Scott Elliott ^68^

Metadata curation, Samples and logistics, and Visualisation:

Hannah Lowe MSc ^25^

Metadata curation, Sequencing and analysis, and Software and analysis tools:

Dr Anna Price PhD ^11^, Matthew R Crown BSc ^65^, Dr Sara Rey PhD ^74^, Dr Sunando Roy PhD ^96^ and Dr Ben Temperton PhD ^105^

Metadata curation, Sequencing and analysis, and Visualisation:

Dr Sharif Shaaban PhD ^73^ and Dr Andrew R Hesketh PhD ^101^

Project administration, Samples and logistics, and Sequencing and analysis:

Dr Kenneth G Laing PhD^41^, Dr Irene M Monahan PhD ^41^ and Dr Judith Heaney PhD ^95, 96, 34^

Project administration, Samples and logistics, and Visualisation:

Dr Emanuela Pelosi FRCPath ^97^, Siona Silviera MSc ^97^ and Dr Eleri Wilson-Davies MD, FRCPath ^97^

Samples and logistics, Software and analysis tools, and Visualisation:

Dr Helen Fryer PhD ^5^

Sequencing and analysis, Software and analysis tools, and Visualization:

Dr Helen Adams PhD ^4^, Dr Louis du Plessis PhD ^23^, Dr Rob Johnson PhD ^39^, Dr William T Harvey PhD ^53, 42^, Dr Joseph Hughes PhD ^53^, Dr Richard J Orton PhD ^53^, Dr Lewis G Spurgin PhD ^59^, Dr Yann Bourgeois PhD ^81^, Dr Chris Ruis PhD ^102^, Áine O’Toole MSc ^104^, Marina Gourtovaia MSc ^116^ and Dr Theo Sanderson PhD ^116^

Funding acquisition, and Leadership and supervision:

Dr Christophe Fraser PhD ^5^, Dr Jonathan Edgeworth PhD, FRCPath ^12^, Prof Judith Breuer MD ^96, 29^, Dr Stephen L Michell PhD ^105^ and Prof John A Todd PhD ^115^

Funding acquisition, and Project administration:

Michaela John BSc ^10^ and Dr David Buck PhD ^115^

Leadership and supervision, and Metadata curation:

Dr Kavitha Gajee MBBS, FRCPath ^37^ and Dr Gemma L Kay PhD ^75^

Leadership and supervision, and Project administration:

Prof Sharon J Peacock PhD ^20, 70^ and David Heyburn ^74^

Leadership and supervision, and Samples and logistics:

Katie Kitchman BSc ^37^, Prof Alan McNally PhD ^43, 93^, David T Pritchard MSc, CSci ^50^, Dr Samir Dervisevic FRCPath ^58^, Dr Peter Muir PhD ^70^, Dr Esther Robinson PhD ^70, 35^, Dr Barry B Vipond PhD ^70^, Newara A Ramadan MSc, CSci, FIBMS ^78^, Dr Christopher Jeanes MBBS ^90^, Danni Weldon BSc ^116^, Jana Catalan MSc ^118^ and Neil Jones MSc ^118^

Leadership and supervision, and Sequencing and analysis:

Dr Ana da Silva Filipe PhD ^53^, Dr Chris Williams MBBS ^74^, Marc Fuchs BSc ^77^, Dr Julia Miskelly PhD

^77^, Dr Aaron R Jeffries PhD ^105^, Karen Oliver BSc ^116^ and Dr Naomi R Park PhD ^116^

Metadata curation, and Samples and logistics:

Amy Ash BSc ^1^, Cherian Koshy MSc, CSci, FIBMS ^1^, Magdalena Barrow ^7^, Dr Sarah L Buchan PhD ^7^, Dr Anna Mantzouratou PhD ^7^, Dr Gemma Clark PhD ^15^, Dr Christopher W Holmes PhD ^16^, Sharon Campbell MSc ^17^, Thomas Davis MSc ^21^, Ngee Keong Tan MSc ^22^, Dr Julianne R Brown PhD ^29^, Dr Kathryn A Harris PhD ^29, 2^, Stephen P Kidd MSc ^33^, Dr Paul R Grant PhD ^34^, Dr Li Xu-McCrae PhD ^35^, Dr Alison Cox PhD ^38, 63^, Pinglawathee Madona ^38, 63^, Dr Marcus Pond PhD ^38, 63^, Dr Paul A Randell MBBCh ^38, 63^, Karen T Withell FIBMS ^48^, Cheryl Williams MSc ^51^, Dr Clive Graham MD ^60^, Rebecca Denton-Smith BSc ^62^, Emma Swindells BSc ^62^, Robyn Turnbull BSc ^62^, Dr Tim J Sloan PhD ^67^, Dr Andrew Bosworth PhD ^70, 35^, Stephanie Hutchings ^70^, Hannah M Pymont MSc ^70^, Dr

Anna Casey PhD ^76^, Dr Liz Ratcliffe PhD ^76^, Dr Christopher R Jones PhD ^79, 105^, Dr Bridget A Knight PhD ^79, 105^, Dr Tanzina Haque PhD, FRCPath ^80^, Dr Jennifer Hart MRCP ^80^, Dr Dianne Irish-Tavares FRCPath ^80^, Eric Witele MSc ^80^, Craig Mower BA ^86^, Louisa K Watson DipHE ^86^, Jennifer Collins BSc ^89^, Gary Eltringham BSc ^89^, Dorian Crudgington ^98^, Ben Macklin ^98^, Prof Miren Iturriza-Gomara PhD ^107^, Dr Anita O Lucaci PhD ^107^ and Dr Patrick C McClure PhD ^113^

Metadata curation, and Sequencing and analysis:

Matthew Carlile BSc ^18^, Dr Nadine Holmes PhD ^18^, Dr Christopher Moore PhD ^18^, Dr Nathaniel Storey PhD ^29^, Dr Stefan Rooke PhD ^73^, Dr Gonzalo Yebra PhD ^73^, Dr Noel Craine DPhil ^74^, Malorie Perry MSc ^74^, Dr Nabil-Fareed Alikhan PhD ^75^, Dr Stephen Bridgett PhD ^77^, Kate F Cook MScR ^84^, Christopher Fearn MSc ^84^, Dr Salman Goudarzi PhD ^84^, Prof Ronan A Lyons MD ^88^, Dr Thomas Williams MD ^104^, Dr Sam T Haldenby PhD ^107^, Jillian Durham BSc ^116^ and Dr Steven Leonard PhD ^116^

Metadata curation, and Software and analysis tools:

Robert M Davies MA (Cantab) ^116^

Project administration, and Samples and logistics:

Dr Rahul Batra MD ^12^, Beth Blane BSc ^20^, Dr Moira J Spyer PhD ^30, 95, 96^, Perminder Smith MSc ^32, 112^, Mehmet Yavus ^85, 109^, Dr Rachel J Williams PhD ^96^, Dr Adhyana IK Mahanama MD ^97^, Dr Buddhini Samaraweera MD ^97^, Sophia T Girgis MSc ^102^, Samantha E Hansford CSci ^109^, Dr Angie Green PhD ^115^, Dr Charlotte Beaver PhD ^116^, Katherine L Bellis ^116, 102^, Matthew J Dorman ^116^, Sally Kay ^116^, Liam Prestwood ^116^ and Dr Shavanthi Rajatileka PhD ^116^

Project administration, and Sequencing and analysis:

Dr Joshua Quick PhD ^43^

Project administration, and Software and analysis tools:

Radoslaw Poplawski BSc ^43^

Samples and logistics, and Sequencing and analysis:

Dr Nicola Reynolds PhD ^8^, Andrew Mack MPhil ^11^, Dr Arthur Morriss PhD ^11^, Thomas Whalley BSc ^11^, Bindi Patel BSc ^12^, Dr Iliana Georgana PhD ^24^, Dr Myra Hosmillo PhD ^24^, Malte L Pinckert MPhil ^24^, Dr Joanne Stockton PhD ^43^, Dr John H Henderson PhD ^65^, Amy Hollis HND ^65^, Dr William Stanley PhD ^65^, Dr Wen C Yew PhD ^65^, Dr Richard Myers PhD ^72^, Dr Alicia Thornton PhD ^72^, Alexander Adams BSc ^74^, Tara Annett BSc ^74^, Dr Hibo Asad PhD ^74^, Alec Birchley MSc ^74^, Jason Coombes BSc ^74^, Johnathan M Evans MSc ^74^, Laia Fina ^74^, Bree Gatica-Wilcox MPhil ^74^, Lauren Gilbert ^74^, Lee Graham BSc ^74^, Jessica Hey BSc ^74^, Ember Hilvers MPH ^74^, Sophie Jones MSc ^74^, Hannah Jones

^74^, Sara Kumziene-Summerhayes MSc ^74^, Dr Caoimhe McKerr PhD ^74^, Jessica Powell BSc ^74^,

Georgia Pugh ^74^, Sarah Taylor ^74^, Alexander J Trotter MRes ^75^, Charlotte A Williams BSc ^96^, Leanne M Kermack MSc ^102^, Benjamin H Foulkes MSc ^109^, Marta Gallis MSc ^109^, Hailey R Hornsby MSc ^109^,

Stavroula F Louka MSc ^109^, Dr Manoj Pohare PhD ^109^, Paige Wolverson MSc ^109^, Peijun Zhang MSc ^109^, George MacIntyre-Cockett BSc ^115^, Amy Trebes MSc ^115^, Dr Robin J Moll PhD ^116^, Lynne Ferguson MSc ^117^, Dr Emily J Goldstein PhD ^117^, Dr Alasdair Maclean PhD ^117^ and Dr Rachael Tomb PhD ^117^

Samples and logistics, and Software and analysis tools:

Dr Igor Starinskij MSc, MRCP ^53^

Sequencing and analysis, and Software and analysis tools:

Laura Thomson BSc ^5^, Joel Southgate MSc ^11, 74^, Dr Moritz UG Kraemer DPhil ^23^, Dr Jayna Raghwani PhD ^23^, Dr Alex E Zarebski PhD ^23^, Olivia Boyd MSc ^39^, Lily Geidelberg MSc ^39^, Dr Chris J Illingworth PhD ^52^, Dr Chris Jackson PhD ^52^, Dr David Pascall PhD ^52^, Dr Sreenu Vattipally PhD ^53^, Timothy M Freeman MPhil ^109^, Dr Sharon N Hsu PhD ^109^, Dr Benjamin B Lindsey MRCP ^109^, Dr Keith James PhD ^116^, Kevin Lewis ^116^, Gerry Tonkin-Hill ^116^ and Dr Jaime M Tovar-Corona PhD ^116^

Sequencing and analysis, and Visualisation:

MacGregor Cox MSci ^20^

Software and analysis tools, and Visualisation:

Dr Khalil Abudahab PhD ^14, 116^, Mirko Menegazzo ^14^, Ben EW Taylor MEng ^14, 116^, Dr Corin A Yeats PhD ^14^, Afrida Mukaddas BTech ^53^, Derek W Wright MSc ^53^, Dr Leonardo de Oliveira Martins PhD ^75^, Dr Rachel Colquhoun DPhil ^104^, Verity Hill ^104^, Dr Ben Jackson PhD ^104^, Dr JT McCrone PhD ^104^, Dr Nathan Medd PhD ^104^, Dr Emily Scher PhD ^104^ and Jon-Paul Keatley ^116^

Leadership and supervision:

Dr Tanya Curran PhD ^3^, Dr Sian Morgan FRCPath ^10^, Prof Patrick Maxwell PhD ^20^, Prof Ken Smith PhD ^20^, Dr Sahar Eldirdiri MBBS, MSc, FRCPath ^21^, Anita Kenyon MSc ^21^, Prof Alison H Holmes MD ^38, 57^, Dr James R Price PhD ^38, 57^, Dr Tim Wyatt PhD ^69^, Dr Alison E Mather PhD ^75^, Dr Timofey Skvortsov PhD ^77^ and Prof John A Hartley PhD ^96^

Metadata curation:

Prof Martyn Guest PhD ^11^, Dr Christine Kitchen PhD ^11^, Dr Ian Merrick PhD ^11^, Robert Munn BSc ^11^, Dr Beatrice Bertolusso Degree ^33^, Dr Jessica Lynch MBCHB ^33^, Dr Gabrielle Vernet MBBS ^33^, Stuart Kirk MSc ^34^, Dr Elizabeth Wastnedge MD ^56^, Dr Rachael Stanley PhD ^58^, Giles Idle ^64^, Dr Declan T Bradley PhD ^69, 77^, Dr Jennifer Poyner MD ^79^ and Matilde Mori BSc ^110^

Project administration:

Owen Jones BSc ^11^, Victoria Wright BSc ^18^, Ellena Brooks MA ^20^, Carol M Churcher BSc ^20^, Mireille Fragakis HND ^20^, Dr Katerina Galai PhD ^20, 70^, Dr Andrew Jermy PhD ^20^, Sarah Judges BA ^20^, Georgina M McManus BSc ^20^, Kim S Smith ^20^, Dr Elaine Westwick PhD ^20^, Dr Stephen W Attwood

PhD ^23^, Dr Frances Bolt PhD ^38, 57^, Dr Alisha Davies PhD ^74^, Elen De Lacy MPH ^74^, Fatima Downing ^74^, Sue Edwards ^74^, Lizzie Meadows MA ^75^, Sarah Jeremiah MSc ^97^, Dr Nikki Smith PhD ^109^ and Luke Foulser ^116^

Samples and logistics:

Dr Themoula Charalampous PhD ^12, 46^, Amita Patel BSc ^12^, Dr Louise Berry PhD ^15^, Dr Tim Boswell PhD ^15^, Dr Vicki M Fleming PhD ^15^, Dr Hannah C Howson-Wells PhD ^15^, Dr Amelia Joseph PhD ^15^, Manjinder Khakh ^15^, Dr Michelle M Lister PhD ^15^, Paul W Bird MSc, MRes ^16^, Karlie Fallon ^16^, Thomas Helmer ^16^, Dr Claire L McMurray PhD ^16^, Mina Odedra BSc ^16^, Jessica Shaw BSc ^16^, Dr Julian W Tang PhD ^16^, Nicholas J Willford MSc ^16^, Victoria Blakey BSc ^17^, Dr Veena Raviprakash MD ^17^, Nicola Sheriff BSc ^17^, Lesley-Anne Williams BSc ^17^, Theresa Feltwell MSc ^20^, Dr Luke Bedford PhD ^26^, Dr James S Cargill PhD ^27^, Warwick Hughes MSc ^27^, Dr Jonathan Moore MD ^28^, Susanne Stonehouse BSc ^28^, Laura Atkinson MSc ^29^, Jack CD Lee MSc ^29^, Dr Divya Shah PhD ^29^, Adela Alcolea-Medina Clinical scientist ^32, 112^, Natasha Ohemeng-Kumi MSc ^32, 112^, John Ramble MSc ^32,112^, Jasveen Sehmi MSc ^32, 112^, Dr Rebecca Williams BMBS ^33^, Wendy Chatterton MSc ^34^, Monika Pusok MSc ^34^, William Everson MSc ^37^, Anibolina Castigador IBMS HCPC ^44^, Emily Macnaughton FRCPath ^44^, Dr Kate El Bouzidi MRCP ^45^, Dr Temi Lampejo FRCPath ^45^, Dr Malur Sudhanva FRCPath ^45^, Cassie Breen BSc ^47^, Dr Graciela Sluga MD, MSc ^48^, Dr Shazaad SY Ahmad MSc ^49, 70^, Dr Ryan P George PhD ^49^, Dr Nicholas W Machin MSc ^49, 70^, Debbie Binns BSc ^50^, Victoria James BSc ^50^, Dr Rachel Blacow MBCHB ^55^, Dr Lindsay Coupland PhD ^58^, Dr Louise Smith PhD ^59^, Dr Edward Barton MD ^60^, Debra Padgett BSc ^60^, Garren Scott BSc ^60^, Dr Aidan Cross MBCHB ^61^, Dr Mariyam Mirfenderesky FRCPath ^61^, Jane Greenaway MSc ^62^, Kevin Cole ^64^, Phillip Clarke ^67^, Nichola Duckworth ^67^, Sarah Walsh ^67^, Kelly Bicknell ^68^, Robert Impey MSc ^68^, Dr Sarah Wyllie PhD^68^, Richard Hopes ^70^, Dr Chloe Bishop PhD ^72^, Dr Vicki Chalker PhD ^72^, Dr Ian Harrison PhD ^72^, Laura Gifford MSc ^74^, Dr Zoltan Molnar PhD ^77^, Dr Cressida Auckland FRCPath ^79^, Dr Cariad Evans PhD ^85, 109^, Dr Kate Johnson PhD ^85, 109^, Dr David G Partridge FRCP, FRCPath ^85, 109^, Dr Mohammad Raza PhD ^85, 109^, Paul Baker MD ^86^, Prof Stephen Bonner PhD ^86^, Sarah Essex ^86^, Leanne J Murray ^86^, Andrew I Lawton MSc ^87^, Dr Shirelle Burton-Fanning MD ^89^, Dr Brendan AI Payne MD ^89^, Dr Sheila Waugh MD ^89^, Andrea N Gomes MSc ^91^, Maimuna Kimuli MSc ^91^, Darren R Murray MSc ^91^, Paula Ashfield MSc ^92^, Dr Donald Dobie MBCHB ^92^, Dr Fiona Ashford PhD ^93^, Dr Angus Best PhD ^93^, Dr Liam Crawford PhD ^93^, Dr Nicola Cumley PhD ^93^, Dr Megan Mayhew PhD ^93^, Dr Oliver Megram PhD ^93^, Dr Jeremy Mirza PhD ^93^, Dr Emma Moles-Garcia PhD ^93^, Dr Benita Percival PhD ^93^, Megan Driscoll BSc ^96^, Leah Ensell BSc ^96^, Dr Helen L Lowe PhD ^96^, Laurentiu Maftei BSc ^96^, Matteo Mondani MSc ^96^, Nicola J Chaloner BSc ^99^, Benjamin J Cogger BSc ^99^, Lisa J Easton MSc^99^, Hannah Huckson BSc ^99^, Jonathan Lewis MSc, PgD, FIBMS ^99^, Sarah Lowdon BSc ^99^, Cassandra S Malone MSc ^99^, Florence Munemo BSc ^99^, Manasa Mutingwende MSc ^99^, Roberto Nicodemi BSc ^99^, Olga Podplomyk FD ^99^, Thomas Somassa BSc ^99^, Dr Andrew Beggs PhD ^100^, Dr Alex Richter PhD ^100^, Claire Cormie ^102^, Joana Dias MSc ^102^, Sally Forrest BSc ^102^, Dr Ellen E Higginson PhD ^102^, Mailis Maes MPhil ^102^, Jamie Young BSc ^102^, Dr Rose K Davidson PhD ^103^, Kathryn A Jackson MSc ^107^, Dr Lance Turtle PhD, MRCP ^107^, Dr Alexander J Keeley MRCP ^109^, Prof Jonathan Ball PhD ^113^, Timothy Byaruhanga MSc ^113^, Dr Joseph G Chappell PhD ^113^, Jayasree Dey MSc ^113^, Jack D Hill MSc ^113^, Emily J Park MSc ^113^, Arezou Fanaie MSc ^114^, Rachel A Hilson MSc ^114^, Geraldine Yaze MSc ^114^ and Stephanie Lo ^116^

Sequencing and analysis:

Safiah Afifi BSc ^10^, Robert Beer BSc ^10^, Joshua Maksimovic FD ^10^, Kathryn McCluggage Masters ^10^, Karla Spellman FD ^10^, Catherine Bresner BSc ^11^, William Fuller BSc ^11^, Dr Angela Marchbank BSc ^11^, Trudy Workman HNC ^11^, Dr Ekaterina Shelest PhD ^13, 81^, Dr Johnny Debebe PhD ^18^, Dr Fei Sang PhD ^18^, Dr Marina Escalera Zamudio PhD ^23^, Dr Sarah Francois PhD ^23^, Bernardo Gutierrez MSc ^23^, Dr Tetyana I Vasylyeva DPhil ^23^, Dr Flavia Flaviani PhD ^31^, Dr Manon Ragonnet-Cronin PhD ^39^, Dr Katherine L Smollett PhD ^42^, Alice Broos BSc ^53^, Daniel Mair BSc ^53^, Jenna Nichols BSc ^53^, Dr Kyriaki Nomikou PhD ^53^, Dr Lily Tong PhD ^53^, Ioulia Tsatsani MSc ^53^, Prof Sarah O’Brien PhD ^54^, Prof Steven Rushton PhD ^54^, Dr Roy Sanderson PhD ^54^, Dr Jon Perkins MBCHB ^55^, Seb Cotton MSc ^56^, Abbie Gallagher BSc ^56^, Dr Elias Allara MD, PhD ^70, 102^, Clare Pearson MSc ^70, 102^, Dr David Bibby PhD ^72^, Dr Gavin Dabrera PhD ^72^, Dr Nicholas Ellaby PhD ^72^, Dr Eileen Gallagher PhD ^72^, Dr Jonathan Hubb PhD ^72^, Dr Angie Lackenby PhD ^72^, Dr David Lee PhD ^72^, Nikos Manesis ^72^, Dr Tamyo Mbisa PhD ^72^, Dr Steven Platt PhD ^72^, Katherine A Twohig ^72^, Dr Mari Morgan PhD ^74^, Alp Aydin MSci ^75^, David J Baker BEng ^75^, Dr Ebenezer Foster-Nyarko PhD ^75^, Dr Sophie J Prosolek PhD ^75^, Steven Rudder ^75^, Chris Baxter BSc ^77^, Sílvia F Carvalho MSc ^77^, Dr Deborah Lavin PhD ^77^, Dr Arun Mariappan PhD ^77^, Dr Clara Radulescu PhD ^77^, Dr Aditi Singh PhD ^77^, Miao Tang MD ^77^, Helen Morcrette BSc ^79^, Nadua Bayzid BSc ^96^, Marius Cotic MSc ^96^, Dr Carlos E Balcazar PhD ^104^, Dr Michael D Gallagher PhD ^104^, Dr Daniel Maloney PhD ^104^, Thomas D Stanton BSc ^104^, Dr Kathleen A Williamson PhD ^104^, Dr Robin Manley PhD ^105^, Michelle L Michelsen BSc ^105^, Dr Christine M Sambles PhD ^105^, Dr David J Studholme PhD ^105^, Joanna Warwick-Dugdale BSc ^105^, Richard Eccles MSc ^107^, Matthew Gemmell MSc ^107^, Dr Richard Gregory PhD ^107^, Dr Margaret Hughes PhD ^107^, Charlotte Nelson MSc ^107^, Dr Lucille Rainbow PhD ^107^, Dr Edith E Vamos PhD ^107^, Hermione J Webster BSc ^107^, Dr Mark Whitehead PhD ^107^, Claudia Wierzbicki BSc ^107^, Dr Adrienn Angyal PhD ^109^, Dr Luke R Green PhD ^109^, Dr Max Whiteley PhD ^109^, Emma Betteridge BSc ^116^, Dr Iraad F Bronner PhD ^116^, Ben W Farr BSc ^116^, Scott Goodwin MSc ^116^, Dr Stefanie V Lensing PhD ^116^, Shane A McCarthy ^116, 102^, Dr Michael A Quail PhD ^116^, Diana Rajan MSc ^116^, Dr Nicholas M Redshaw PhD ^116^, Carol Scott ^116^, Lesley Shirley MSc ^116^ and Scott AJ Thurston BSc ^116^

Software and analysis tools:

Dr Will Rowe PhD^43^, Amy Gaskin MSc ^74^, Dr Thanh Le-Viet PhD ^75^, James Bonfield BSc ^116^, Jennifier Liddle ^116^ and Andrew Whitwham BSc ^116^

1 Barking, Havering and Redbridge University Hospitals NHS Trust, 2 Barts Health NHS Trust, 3 Belfast Health & Social Care Trust, 4 Betsi Cadwaladr University Health Board, 5 Big Data Institute, Nuffield Department of Medicine, University of Oxford, 6 Blackpool Teaching Hospitals NHS Foundation Trust, 7 Bournemouth University, 8 Cambridge Stem Cell Institute, University of Cambridge, 9 Cambridge University Hospitals NHS Foundation Trust, 10 Cardiff and Vale University Health Board, 11 Cardiff University, 12 Centre for Clinical Infection and Diagnostics Research, Department of Infectious Diseases, Guy’s and St Thomas’ NHS Foundation Trust, 13 Centre for Enzyme Innovation, University of Portsmouth, 14 Centre for Genomic Pathogen Surveillance, University of Oxford, 15 Clinical Microbiology Department, Queens Medical Centre, Nottingham University Hospitals NHS Trust, 16 Clinical Microbiology, University Hospitals of Leicester NHS Trust, 17 County Durham and Darlington NHS Foundation Trust, 18 Deep Seq, School of Life Sciences, Queens Medical Centre, University of Nottingham, 19 Department of Infectious Diseases and Microbiology, Cambridge University Hospitals NHS Foundation Trust, 20 Department of Medicine, University of Cambridge, 21 Department of Microbiology, Kettering General Hospital, 22 Department of Microbiology, South West London Pathology, 23 Department of Zoology, University of Oxford, 24 Division of Virology, Department of Pathology, University of Cambridge, 25 East Kent Hospitals University NHS Foundation Trust, 26 East Suffolk and North Essex NHS Foundation Trust, 27 East Sussex Healthcare NHS Trust, 28 Gateshead Health NHS Foundation Trust, 29 Great Ormond Street Hospital for Children NHS Foundation Trust, 30 Great Ormond Street Institute of Child Health (GOS ICH), University College London (UCL), 31 Guy’s and St. Thomas’ Biomedical Research Centre, 32 Guy’s and St. Thomas’ NHS Foundation Trust, 33 Hampshire Hospitals NHS Foundation Trust, 34 Health Services Laboratories, 35 Heartlands Hospital, Birmingham, 36 Hub for Biotechnology in the Built Environment, Northumbria University, 37 Hull University Teaching Hospitals NHS Trust, 38 Imperial College Healthcare NHS Trust, 39 Imperial College London, 40 Infection Care Group, St George’s University Hospitals NHS Foundation Trust, 41 Institute for Infection and Immunity, St George’s University of London, 42 Institute of Biodiversity, Animal Health & Comparative Medicine, 43 Institute of Microbiology and Infection, University of Birmingham, 44 Isle of Wight NHS Trust, 45 King’s College Hospital NHS Foundation Trust, 46 King’s College London, 47 Liverpool Clinical Laboratories, 48 Maidstone and Tunbridge Wells NHS Trust, 49 Manchester University NHS Foundation Trust, 50 Microbiology Department, Buckinghamshire Healthcare NHS Trust, 51 Microbiology, Royal Oldham Hospital, 52 MRC Biostatistics Unit, University of Cambridge, 53 MRC-University of Glasgow Centre for Virus Research, 54 Newcastle University, 55 NHS Greater Glasgow and Clyde, 56 NHS Lothian, 57 NIHR Health Protection Research Unit in HCAI and AMR, Imperial College London, 58 Norfolk and Norwich University Hospitals NHS Foundation Trust, 59 Norfolk County Council, 60 North Cumbria Integrated Care NHS Foundation Trust, 61 North Middlesex University Hospital NHS Trust, 62 North Tees and Hartlepool NHS Foundation Trust, 63 North West London Pathology, 64 Northumbria Healthcare NHS Foundation Trust, 65 Northumbria University, 66 NU-OMICS, Northumbria University, 67 Path Links, Northern Lincolnshire and Goole NHS Foundation Trust, 68 Portsmouth Hospitals University NHS Trust, 69 Public Health Agency, Northern Ireland, 70 Public Health England, 71 Public Health England, Cambridge, 72 Public Health England, Colindale, 73 Public Health Scotland, 74 Public Health Wales, 75 Quadram Institute Bioscience, 76 Queen Elizabeth Hospital, Birmingham, 77 Queen’s University Belfast, 78 Royal Brompton and Harefield Hospitals, 79 Royal Devon and Exeter NHS Foundation Trust, 80 Royal Free London NHS Foundation Trust, 81 School of Biological Sciences, University of Portsmouth, 82 School of Health Sciences, University of Southampton, 83 School of Medicine, University of Southampton, 84 School of Pharmacy & Biomedical Sciences, University of Portsmouth, 85 Sheffield Teaching Hospitals NHS Foundation Trust, 86 South Tees Hospitals NHS Foundation Trust, 87 Southwest Pathology Services, 88 Swansea University, 89 The Newcastle upon Tyne Hospitals NHS Foundation Trust, 90 The Queen Elizabeth Hospital King’s Lynn NHS Foundation Trust, 91 The Royal Marsden NHS Foundation Trust, 92 The Royal Wolverhampton NHS Trust, 93 Turnkey Laboratory, University of Birmingham, 94 University College London Division of Infection and Immunity, 95 University College London Hospital Advanced Pathogen Diagnostics Unit, 96 University College London Hospitals NHS Foundation Trust, 97 University Hospital Southampton NHS Foundation Trust, 98 University Hospitals Dorset NHS Foundation Trust, 99 University Hospitals Sussex NHS Foundation Trust, 100 University of Birmingham, 101 University of Brighton, 102 University of Cambridge, 103 University of East Anglia, 104 University of Edinburgh, 105 University of Exeter, 106 University of Kent, 107 University of Liverpool, 108 University of Oxford, 109 University of Sheffield, 110 University of Southampton, 111 University of St Andrews, 112 Viapath, Guy’s and St Thomas’ NHS Foundation Trust, and King’s College Hospital NHS Foundation Trust, 113 Virology, School of Life Sciences, Queens Medical Centre, University of Nottingham, 114 Watford General Hospital, 115 Wellcome Centre for Human Genetics, Nuffield Department of Medicine, University of Oxford, 116 Wellcome Sanger Institute, 117 West of Scotland Specialist Virology Centre, NHS Greater Glasgow and Clyde, 118 Whittington Health NHS Trust

COVID-19 Impact Project (Trinidad & Tobago Group)

Funding Acquisition, Leadership and Supervision, Study Conception, Design and Administration:

Prof. Christine V. F. Carrington ^1^

Study Design, Sequencing and Analysis, Logistics, Data curation, Database:

Dr. Nikita Sahadeo ^1^

Sequencing and Analysis:

Vernie Ramkissoon ^1^

Study Design, Sequencing and Analysis:

Dr. Sarah Hill ^2,3^

Study Design and Supervision:

Prof. Christopher Oura ^4^

Study Design, Samples and Logistics Dr. Gabriel Gonzales Escobar ^5^

Samples and Logistics:

Dr. Arianne Brown-Jordan ^6^, SueMin Nathaniel Girdharrie ^5^, Risha Singh ^5^, Dr. Avery Q J Hinds ^6^, Dr. Naresh Nandram ^6^, Dr. Roshan Parasram ^6^, Lisa Edghill ^5^, Dr. Lisa Indar ^5^, Zobida Khan-Mohammed ^6^, Dr. Joy St. John ^5^

Data Curation and Database:

Anushka Ramjag ^1^

Study Design:

Prof. Oliver G. Pybus ^2^, Dr. Nuno Faria ^2,7^, Dr. Jerome Foster ^1^

1 Department of Preclinical Sciences, Faculty of Medical Sciences, The University of the West Indies, St. Augustine, Trinidad and Tobago, 2 Department of Zoology, University of Oxford, Oxford, UK, 3 Royal Veterinary College, University of London, UK, 4 Department of Basic Veterinary Sciences, Faculty of Medical Sciences, The University of the West Indies, St. Augustine, Trinidad and Tobago, 5 Caribbean Public Health Agency, Port-of-Spain, Trinidad and Tobago, 6 Ministry of Health, Port-of-Spain, Trinidad and Tobago, 7 Department of Infectious Disease Epidemiology, Imperial College, London, UK.

